# The Effect of Postnatal Care on the Postpartum Contraceptive Use in Ethiopia: A Systematic Review and Meta-analysis

**DOI:** 10.1101/2023.05.23.23290396

**Authors:** Tesfalem Tilahun Yemane, Mengestu Mera, Abebe Kassa, Nigusie Tadesse

## Abstract

**Background:** Postpartum contraceptive use is an essential aspect of maternal and child health. The use of contraception after childbirth is a critical step in ensuring healthy spacing between births, reducing maternal and infant mortality. However, it is often observed that the use of postpartum contraception is low. In this context, postnatal care (PNC) visits are an important opportunity to promote postpartum contraceptive use. Therefore, the aim of this review is to examine the effect of postnatal visits on the utilization of postpartum contraceptive use.

**Methods:** We conducted a systematic review and meta-analysis of published and unpublished studies. Pub Med, HINARI, Science direct, Cochrane Library, ETH Library and Google Scholar were used to search all articles. For data analysis, STATA 14 software was used. Funnel plots and Egger’s test were used to examine the risk of publication bias. Heterogeneity was checked by using Cochran’s-Q test and I^2^ test. Random effect model was computed to estimate the pooled prevalence.

**Results:** The finding of the present systematic review and meta-analysis indicated that having postnatal care visits increase the utilization of postpartum modern contraceptive [pooled effect size 2.92 (95% CI, 2.21, 3.881)]. Postnatal care can provide critical information and support to women during the postpartum period, including information about family planning and contraceptive options.

**Conclusion:** The utilization of postpartum contraception is a critical aspect of maternal and child health. Postnatal care visits provide an opportunity to promote postpartum contraceptive use. During these visits, healthcare providers can provide information on the available contraceptive methods, counsel women on the benefits and risks of each method, and assist in the selection of a suitable method.

**PROSPERO Registration Number:** CRD42020156574

## Introduction

Postpartum contraceptive means the prevention of unintended and closely spaced pregnancies during the first 12 months following childbirth [1]. The use of contraception after childbirth is a critical step in ensuring healthy spacing between births, reducing maternal and infant mortality, and promoting family planning [2, 3]. In Ethiopia, the World Health Organization (WHO) and Ethiopian family planning program recommend an interval of at least 2 years [4, 5] and women recommend receiving postpartum family planning counseling during antenatal, immediate postpartum, and postnatal services [5, 6]. Postnatal care (PNC) presents an opportune moment when women should be counseled on family planning. Contraceptive method options should be discussed and provided during postnatal care [1, 6]. Short birth intervals increase the risk for the health of both the mother and the child, such as the risk of preterm birth, low birth weight and small for gestational age, increased chances of chronic undernourishment, stunted growth, and child mortality [4, 7]. Using family planning during the postpartum period may help women to space births by at least 24 months, and this fact can also help to reduce maternal and child mortality by 30% and 10 % respectively [8].

Maternal health remains a major global concern since pregnancy and childbirth are the leading causes of morbidity, mortality, and disability among women of the reproductive age group [9]. Integrating PPFP into MNCH programs and services contributes to the reduction of high-risk pregnancies, reduced unmet need for Family planning, and improvements in the health and survival of mothers and children [10].

Globally, more than 9 out of 10 women want to avoid their pregnancy at least for the next 24 months after childbirth [11]. According to DHS data from 21 low and middle-income countries between 2005 and 2012, sixty-one percent (61%) of postpartum women had an unmet need for family planning [2, 12]. Studies done in five low-income countries showed that postpartum modern contraceptive use and unmet need varied widely. Half of women have an unmet need for family planning services among all women who wants to delay the next pregnancy [13].

Despite global and national efforts, the uptake of postpartum contraceptives in Ethiopia is still low and the unmet need is high; in 2016, it was estimated that 26% of postpartum women were using family planning in the first year of the postpartum period [6, 14]. Since the uptake of PPFP was low, PNC services were a significant variable, it creates windows of opportunity to provide family planning counseling and it leads to offering women modern contraceptive methods [6, 15-17].

Postnatal care visits are a crucial opportunity to promote postpartum contraceptive use [6, 15, 18]. However, the effectiveness of postnatal visits in promoting postpartum contraceptive use is not well understood. Therefore, the aim of this systematic review and meta-analysis is to analyze and summarize the effect of postnatal care services on the uptake of postpartum family planning in Ethiopia. This evidence will help the government of Ethiopia and other stakeholders who work in family planning.

## Materials and Methods

### Study Design and Protocol Registration

A systematic review and meta-analysis were conducted to quantify the pooled effect of PNC on postpartum contraceptive use. To report this systematic review and meta-analysis, an updated Preferred Reporting Items for Systematic Review and Meta-Analysis (PRISMA) 2020 statement was adapted [19]. The protocol has been registered on the International Prospective Register of Systematic Reviews, the University of York Center for Reviews and Dissemination. Available from: https://www.crd.york.ac.uk/prospero/display_record.php? (Registration ID number =CRD42020156574).

#### Search strategies

PubMed, HINARI, Science Direct, Cochrane Library electronic databases, Google Search and Google Scholar were systematically searched. Medical subject heading (MeSH), keywords, and gloss were used to identify the components of PICO. Boolean operators (“OR”, ‘‘AND” and “NOT”) were used to combine different search terms. The following keywords were used in the title search: “Postpartum family planning” OR “Postpartum contraceptive” AND Prevalence OR Epidemiology OR utilization OR use AND determinants OR déterminante OR “Factors associated” OR predictors OR postnatal OR ‘postnatal care’. This systematic review and meta-analysis used the PICO (Population, Intervention, Comparison, and Outcomes) framework to identify the eligible articles. The study Population (P) were reproductive age (15-49 years) group women in their first 12 months after delivery, the Intervention (I) was PNC follow-up, the Comparison (C) group were women who did not have PNC follow-up, and the Outcomes (O) of this study were the utilization of postpartum contraceptive within 12 months after delivery.

### Criteria for eligibility

#### Inclusion criteria

All studies reported in the English language, published and unpublished studies, and articles that revealed the association between postnatal care visits and postpartum contraceptive was included in this review. Studies conducted in Ethiopia and published between January 30/ 2013 and January 30/2023 were included.

#### Exclusion Criteria

Studies that did not report the PNC visit status as well as their outcomes were excluded. In addition, articles that were not fully accessed after at least two email contacts of the primary author or failed to contact their primary authors were excluded.

#### Study selection

All searched articles for the review were imported to EndNote X7 and duplicated studies were excluded. All studies were initially examined for inclusion based on information contained in the titles alone and abstracts, and then a full-text review was performed by two independent reviewers (TTY and AK). Potentially eligible articles were identified by two independent reviewers (TTY and AK). Disagreements between the reviewers (TTY and AK) were solved by discussion & additional reviewer (NT). The PRISMA flow diagrams were used to summarize the overall selection process of the article [19].

### Quality assessment of the studies

Three reviewers (TTY, NT, and MM) independently assessed the quality of the included studies. A modified version of the Newcastle-Ottawa Scales was used to evaluate the quality of the studies [20]. The studies were divided into three categories: (0–4) low quality, (5–7) medium quality, and (8–10) high quality [21]. Those studies with medium (satisfying 50%) and high quality were included for analysis.

### Data extraction

All data were extracted by two independent reviewers (TTY and MM). The discrepancies were solved through discussion & involving the third reviewer (AK). From each included article, the name of the author/s, publication year, study area & region, study setting, study design, sample size, and adjusted odds ratio with confidence interval were extracted. The extracted data were entered into a Microsoft Excel spreadsheet before being exported to the STATA 14 software.

### Data analysis, heterogeneity, and Publication bias

STATA Version 14 (software) was used to calculate the pooled effect size with 95% confidence intervals (95% CI). Statistical heterogeneity was checked by using the Cochran’s-Q test and the I^2^ test. The I^2^ statistics of below 25% are low heterogeneity, 25–50% is moderate heterogeneity, 51–75% is substantial heterogeneity, and above 75% is considerable heterogeneity [22]. A p-value <0.05 was used to declare heterogeneity. Visual inspection of the funnel plots, Egger’s weighted regression, and Begg’s rank correlation tests were used to examine the possible risk of publication bias. Forest plots and Odds Ratios with their 95% CI were used to present the pooled effect sizes. We use a random effects model was used to estimate the pooled effect due to the presence of heterogeneity. Subgroup analysis was conducted by region, publication year, study setting, and sample size.

## Results and Discussion

### Study selection

A total of 1,208 articles were searched through electronic databases and gray pieces of literature. From these, 144 articles were excluded due to duplications, while the remaining 1,064 articles were reserved for further screening. Of these remaining articles, 776 and 221 articles were excluded by their titles and abstracts, respectively. A total of 67 full-text articles were assessed for eligibility criteria. Finally, 10 articles with appropriate quality were included in the final systematic review and meta-analysis. Furthermore, to summarize the selection procedure the PRISMA flow diagram was used. (Fig 1)

**Figure 1.**
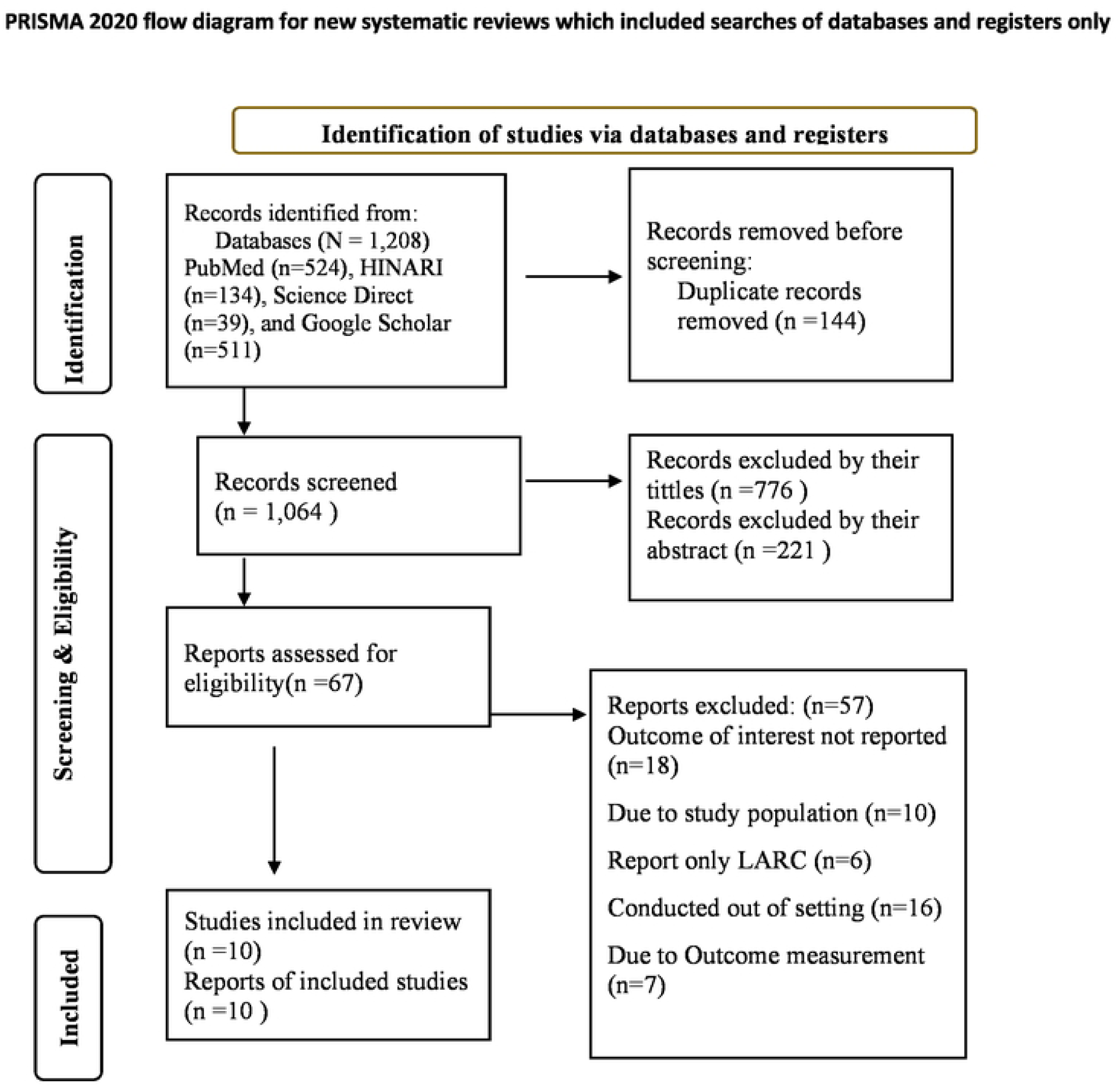
Description of schematic presentation of the PRISMA flow diagram to select and include studies, 2023. *From:* Page MJ, McKenzie JE, Bossuyt PM, Boutron I, Hoffmann TC, Mulrow CD, et al. The PRISMA 2020 statement: an updated guideline for reporting systematic reviews. For more information, visit: http://www.prismastatement.org/

**Figure 2:**
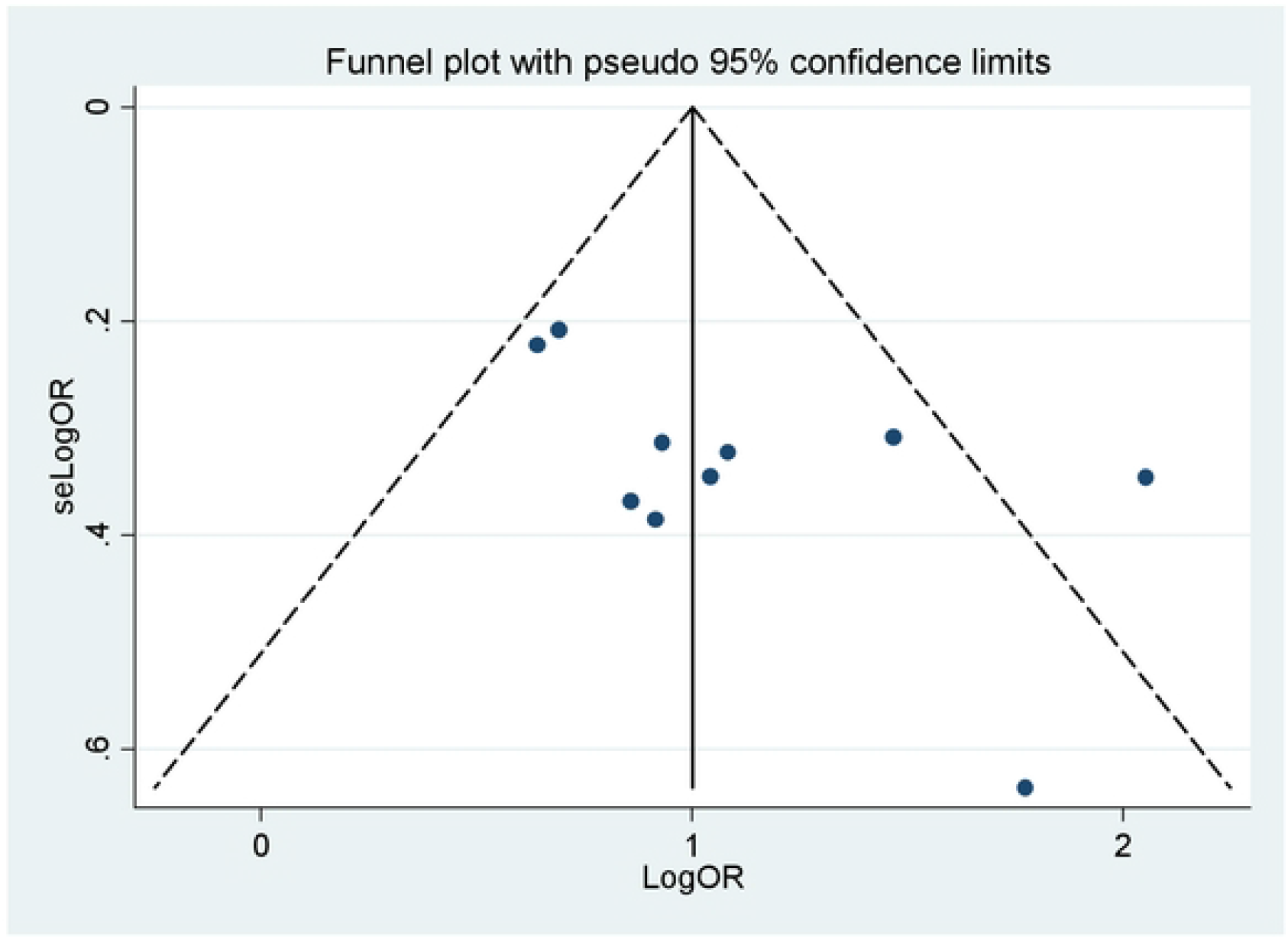
Funnel plot of the 10 included studies of the effect of PNC on utilization of postpartum contraceptives in Ethiopia, 2023. The vertical line indicates the effect size whereas the diagonal line indicates the precision of individual studies with a 95% confidence interval.

**Figure 3:**
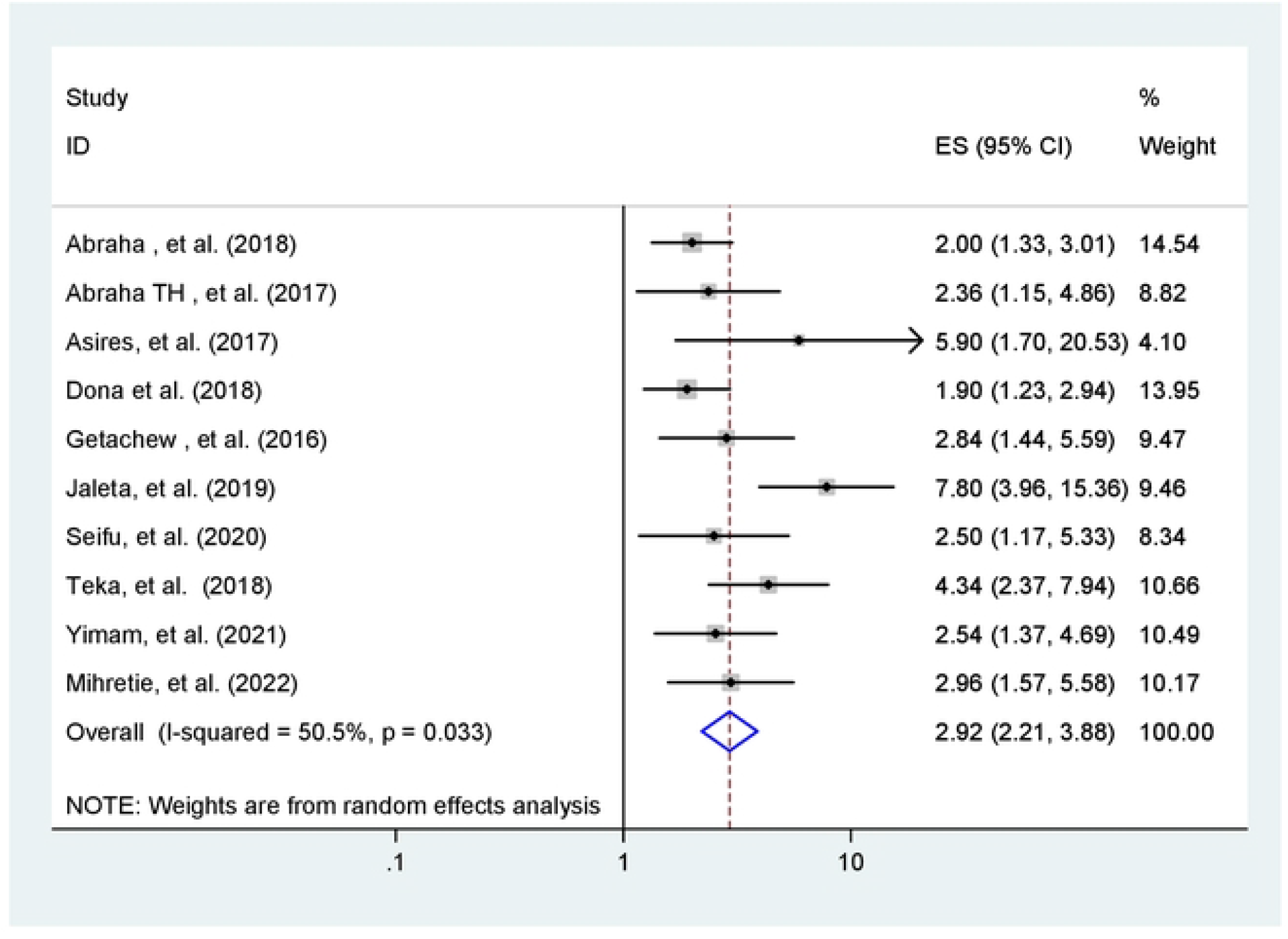
Forest plot of 10 included studies, which reveal the effect of PNC on utilization of postpartum contraceptive in Ethiopia, 2023.

### Characteristics of Included Studies

As described in **Error! Reference source not found**., these, 10 original articles were included in this systematic review and meta-analysis. The selected articles were published from 2016 to 2022. One of them was unpublished articles [23]. All included studies are cross-sectional in nature. In this study, 6157 postpartum women were involved. The sample size of the included studies ranged from 342 [17] to 1109 [16]. Out of 10 studies, 6 studies were conducted both in urban and rural settings[15, 18, 23-26], Three in urban[27-29], and one in rural [16] settings. The mean age of postpartum women ranged from 25.4 [24] to 29.82 years[28]. The response rate of included studies ranges from 95.6% [25] to 100 % response rate[16, 27]. Finally, the quality score of the included studies ranges from 7 up to 8 out of 10 points. Further descriptions and characteristics of the studies included in (**Error! Reference source not found**.).

### Quality of the Studies

The quality of the selected articles was assessed by Newcastle-Ottawa Scale modified versions [20]. Regarding the quality score of the included studies, 7 of the 10 studies had high quality (8-10 points) and the remaining 3 studies were medium level quality (5-7 points). Therefore, studies that had considerable risk were not included in this review (Table 1).

### Publication bias and Heterogeneity

Publication bias was presented on traditional funnel plots. The shape of the traditional funnel plots indicates symmetrical distribution (**Error! Reference source not found**.). Moreover, to ascertain the funnel plot, Begg’s and Egger’s tests were conducted. Begg’s and Egger’s test results revealed the absence of statistically significant publication bias (p = 0.28 and p = 0.060, respectively). Statistical heterogeneity was checked by using the Cochran’s-Q test and the I^2^ test and through visual examination of the forest plot (overlap of confidence intervals). In this analysis, moderate heterogeneity was observed across the included studies and detected by the Cochrane Q test (Q test p<0.001) and I^2^ statistics (I^2^ = 50.5%) (**Error! Reference source not found**.). Because of this, a random effects model was used to estimate the effect of PNC follow-up on postpartum contraceptive use. By considering publication year, study setting, and sample size; sub-group analysis was performed to identify the possible source of heterogeneity, but none of these variables were found to be statistically significant (**Error! Reference source not found**.).

### Meta-Analysis

#### The effect of postnatal care follow-up on the utilization of postpartum modern contraceptive

As shown in the forest plot, the results of 10 included studies indicated that the pooled effect size of postpartum modern contraceptive utilization among those mothers who had postnatal care visits was 2.92 (95% CI, 2.21, 3.88) compared to those mothers without having PNC visits in the random effects model (**Error! Reference source not found**.). The finding of this systematic review and meta-analysis revealed that having postnatal care visits increase the utilization of postpartum modern contraceptive.

#### Subgroup analysis

To decrease sizeable heterogeneity, subgroup analysis was performed based on the regions where the studies were conducted, the sample size of the studies, the study setting, and the year of publication. Based on subgroup analysis, the effect of postnatal care follow-up on utilization in the Tigray region was 2.08, 95% CI (1.46, 2.97), Southern Nation and Nationalities 2.14, 95% CI (1.48, 3.08) and Oromia was 3.46, 95% CI (2.03, 5.89) (**Error! Reference source not found**.). With regard to sample size, the higher the sample size the more precise the effect size.

## Discussion

The findings of this systematic review and meta-analysis revealed that PNC follow-up has a significant effect on the utilization of postpartum contraceptives. The current meta-analysis reported that having postnatal care services were positively associated [2.92 (95% CI, 2.21, 3.88)] with the use of contraceptive during the postpartum period. This suggests that women who receive postnatal care are almost three times more likely to use contraceptives after giving birth compared to those who do not receive postnatal care.

This finding is consistent with USAID findings from 17 Countries, a systematic review and meta-analysis conducted in low- and middle-income countries, and low-income countries of SSA [30-32]. Those findings suggest that the utilization of contraceptives among postpartum women will increase substantially if more women use postnatal care.

The possible explanation for this finding is that women might get the opportunity for contraceptive counseling and the benefits of birth spacing from health professionals during PNC follow-up. In addition, PNC provides an important opportunity for healthcare providers to deliver appropriate contraceptive methods.

The techniques for testing the publication bias of included studies were considered. It was examined by performing Egger’s correlation and Begg’s regression intercept tests at a 5% significant level. The results of Begg’s and Egger’s tests indicated that there was no statistically significant publication bias across the included studies as evidenced by p = 0.28 and p = 0.06 in Begg’s and Egger’s tests respectively. In this meta-analysis, to identify the possible sources of heterogeneity, subgroup analysis was performed based on the regions and sample size. However, the result of the subgroup analysis indicated that the source of moderate heterogeneity was not because of the study regions and sample size.

### Limitations of the Study

Only English articles were considered to conduct this review. In addition, the majority of studies were obtained from Oromia, Tigray & SNNP regions. Therefore, the results may not be strongly representative of the other regions due to the small number of studies included.

## Conclusions

In conclusion, this systematic review and meta-analysis revealed that the pooled effect of postnatal care on postpartum contraceptive use in Ethiopia was found to be significant, with women who receive postnatal care being almost three times more likely to use contraceptives after giving birth compared to those who do not receive postnatal care. Integrating family planning counseling & service in all MCH units could be an effective strategy to increase postpartum contraceptive utilization. Therefore, maximizing postnatal care visits helps to improve the utilization of postpartum contraceptives, as well as for the better outcome of maternal and child health. Thus, having postnatal care visits is strongly recommended.

## Data Availability

Data will be available upon reasonable request from the corresponding author.

## Data Availability

Data will be available upon reasonable request to the corresponding author.

## Acknowledgments

Not applicable

## Authors’ contributions

All authors have contributed to this study. TTY contributed to the conception of research protocol, protocol preparation and registration, study design, literature review, data extraction, data analysis, interpretation, and drafting of the manuscript. NT participated in literature review, data extraction, and quality assessment. AK participated in literature review, data extraction, and quality assessment. MM participated in literature review, data extraction, and quality assessment. All authors read and approved the final manuscript.

## Supporting information

S1 Fig 1: Description of schematic presentation of the PRISMA 2020 flow diagram to select and include studies, 2023.

S2 Fig 2: Funnel plot of the 10 included studies of the effect of PNC on utilization of postpartum contraceptives in Ethiopia, 2023.

S2 Fig 3: Forest plot of 10 included studies, which reveal the effect of PNC on utilization of postpartum contraceptive in Ethiopia, 2023.

S1 Table 1: Descriptive summary of 10 included studies in the systematic review and meta-analysis of the effect of postnatal care on postpartum family planning use among women of the reproductive age group in Ethiopia, 2023.

S2 Table 2: Subgroup analysis for the effect of postnatal care on postpartum family planning use among women of the reproductive age (15-49 years) group in their first 12 months after delivery in Ethiopia, 2023 (n = 10).

S1 File: PRISMA 2020 Checklist

